# Supraglottic Airway Devices Improve Clinical Outcomes for Out-of-Hospital Cardiac Arrest: An Updated Systematic Review and Trial Sequential Meta-Analysis Involving 196,573 Patients

**DOI:** 10.64898/2025.12.29.25343184

**Authors:** Li-fu Zheng, Jin-zhou Feng, Ying-Yi Ding, Lei Deng, Jun Zeng, Charles Damien Lu, Ming-wei Sun, Hua Jiang

## Abstract

**OBJECTIVES:** Although several systematic reviews attempted to compare supraglottic airway (SGA) and endotracheal intubation for or adults with out-of-hospital cardiac arrest (OHCA), the optimal airway management strategy remains debated. We conducted a systematic review and meta-analysis comparing supraglottic airway (SGA) devices and endotracheal intubation (ETI) as initial interventions in OHCA to clarify their relative clinical efficacy.

**DATA SOURCES:** We retrieved relevant clinical trials from PubMed, Embase, Cochrane Library, Web of Science, and China National Knowledge Infrastructure(CNKI), as well as unpublished sources, from inception to October 10, 2025.

**STUDY SELECTION:** We included randomized controlled trials (RCTs) and non-randomized studies involving adult patients with OHCA who were assigned to supraglottic airway (SGA) versus endotracheal intubation for initial prehospital airway management.

**RESULTS:** Our systematic review and meta-analysis enrolled 9 randomized controlled trials and 23 non-randomized studies, involving 196,573 patients. Patients who received SGA were associated with higher incidence of return of spontaneous circulation (ROSC), (RCT RR = 1.17, 95% CI: [1.03,1.34], P = 0.016, I² = 48.5%), higher incidence of first-attempt success rates (RCT: RR = 1.31, 95% CI, [1.14–1.51], P = 0.0029. Non-RCTs: RR = 1.46, 95% CI: [1.26,1.70], P < 0.0001, I² = 83.1%), higher incidence of return of time to airway securement (RCT: MD = -2.30 min, 95% CI: [-3.54, -1.05], P = 0.0003, I² = 98.4%. Non-RCTs: MD = -2.86 min, 95% CI: [-3.62, -2.11], P < 0.0001, I² = 95.8%.). There is no difference in favorable neurological outcome (RCT: RR = 1.06, 95% CI: [0.84, 1.35], P = 0.6202, I² = 45.4%. Non-RCTs: RR = 0.94, 95% CI: [ 0.76, 1.18], P = 0.6198, I² = 74.2%.) and no difference in occurrence of regurgitation/aspiration (RR 1.03; 95% CI: [0.93, 1.14]; P=0.5426, I² = 0.0%). Additionally, trial sequential analysis was performed to validate these findings.

**CONCLUSION:** For adult patients experiencing out-of-hospital cardiac arrest (OHCA), initial airway management by using supraglottic airway (SGA) improves rate of return of spontaneous circulation (ROSC), enables faster airway placement, and achieves higher first-pass success rate when compared with endotracheal intubation. There is a high degree of certainty regarding the major outcomes.

## 1 Introduction

Out-of-hospital cardiac arrest (OHCA), defined as the cessation of cardiac activity occurring outside the hospital setting or prior to hospital admission, represents a frequent critical condition encountered in emergency departments [1]. According to the World Health Organization (WHO), cardiac arrest contributes to more than 5 million deaths globally each year [2]. For patients with OHCA, high-quality chest compression and effective ventilation are essential components of successful resuscitation [3]. Currently, the primary ventilation devices employed by Emergency Medical Services (EMS) worldwide include endotracheal intubation (ETI), supraglottic airway devices (SGA), and bag-valve-mask (BVM) ventilation [4]. Nevertheless, the optimal strategy for advanced airway management during cardiac arrest remains a subject of debate [5].

Regardless of the technique selected, advanced airway management must be performed by healthcare professionals once the patient arrives at the emergency department. Endotracheal intubation is widely regarded as the first-line method for securing the airway. Both the 2023 American Heart Association (AHA) guidelines and the European Resuscitation Council (ERC) guidelines recommend endotracheal intubation as the preferred approach for advanced airway management when performed by experienced providers, as it ensures reliable oxygenation and ventilation while reducing the risk of aspiration[6, 7]. When endotracheal intubation fails or when the provider lacks sufficient proficiency, supraglottic airways (such as laryngeal masks or i-gel) may serve as suitable alternatives due to their ease of insertion and minimal interruption to chest compressions[7]. However, several studies indicate that emergency physicians experience relatively high failure rates during first-attempt intubation both in emergency and in-hospital settings[8]. Moreover, successful endotracheal intubation is strongly correlated with operator experience: senior clinicians demonstrate higher first-pass success rates, whereas junior clinicians often encounter challenges due to limited expertise[9].

Supraglottic airway devices (SGA) comprise a class of airway management devices designed to be inserted into the pharynx, enabling ventilation, oxygenation, and delivery of anesthetic gases without entering the trachea. Common examples include laryngeal masks, newer-generation i-gel devices, and laryngeal tubes[10]. Compared to ETI, SGA insertion is generally associated with greater ease of use, with studies reporting higher first-attempt success rates and reduced time to secure the airway [8, 11, 12]. Emerging evidence also suggests that SGAs can achieve ventilatory efficacy comparable to that of endotracheal intubation (ETI) [13, 14].

Therefore, we conducted this systematic review and meta-analysis to synthesize evidence and observational studies comparing SGA insertion and ETI in the prehospital initial airway management of OHCA.

## 2 MATERIALS AND METHODS

### 2.1 Protocol registration and reporting format

This systematic review and meta-analysis followed the recommendations of the Preferred Reporting Items for Systematic Reviews and Meta-Analyses (PRISMA) statement. The protocol for this systematic review has been registered in the PROSPERO database (ID: CRD420250642647).

### 2.2 Data sources and search strategy

We performed our literature retrieval from inception to December 20, 2025. The search covered multiple databases, including MEDLINE (accessed via PubMed), the Cochrane Library, Web of Science, and the China National Knowledge Infrastructure (CNKI). Additionally, we searched trial registries, specifically the Chinese Clinical Trial Registry (www.chictr.org.cn) and ClinicalTrials.gov, for ongoing or completed clinical trials. Our search strategy incorporated the following key terms: “clinical trial”, “out of hospital cardiac arrest”, “supraglottic airway”, and “tracheal intubation”.

We established the following inclusion criteria based on participants, interventions, comparisons, outcomes, and study design (i.e., the PICOS framework):

1. Participants: patients (≥ 18 years old) with OHCA.
2. Intervention: SGA insertion.
3. Comparison: Tracheal intubation
4. Outcome:
5. Major outcomes: ROSC, Time to advanced airway placement, The success rate of the first attempt.
6. Secondary outcome: Survival with good functional outcome at longest follow-up.

Adverse events, such as aspiration.

1. 5) Study design: Randomized controlled trial (RCT) and cohort studies.

Exclusion criteria

1. Historically outdated airway devices, including the esophageal tracheal combitube, pharyngeal tracheal lumen airway, and esophageal obturator airways.
2. Age < 18 years
3. Duplicated literature
4. Animal experiments

### 2.3 Assessment of study quality

Two researchers, Li-Fu Zheng and Ying-Yi Ding, used two methods to assess the literature quality. Randomized controlled trials were evaluated with the modified version of the Cochrane Risk of Bias 2 (RoB 2) tool[15]. We assessed risk of bias (RoB) independently and in duplicate using the modified Cochrane RoB 2 tool, for which each domain is rated as “low,” “probably low,” “high,” or “probably high.” Cohort studies were evaluated with The Risk Of Bias In Non-randomized Studies of Interventions (ROBINS-I) assessment tool[15]. Each domain was rated as "Low risk of bias," "Moderate risk of bias," "Serious risk of bias," or "Critical risk of bias" (following the ROBINS-I tool guidelines).

### 2.4 Statistical method

We employed RevMan 5.4 (The Cochrane Collaboration), Stata (StataMP 18), and R (R Foundation for Statistical Computing 4.1.1) as our primary tools for conducting meta-analyses. For continuous outcome variables, effect sizes were calculated as either the Standardized Mean Difference (SMD) or the Weighted Mean Difference (WMD). For dichotomous outcome variables, pooled effect estimates were expressed as Risk Ratios (RR) with corresponding 95% Confidence Intervals (CI).

To evaluate the risk of type I error (false positive results) in our meta-analyses and to determine the required information size (RIS), we performed Trial Sequential Analysis (TSA). TSA also aids in establishing potential stopping boundaries for future clinical trials, thereby potentially preventing unnecessary resource expenditure by indicating when sufficient evidence has been accumulated.

TSA was conducted using TSA version 0.9.5.10 Beta software (Copenhagen Trial Unit, Centre for Clinical Intervention Research, Denmark). The analysis settings employed a fixed-effect model, with type I (α) and type II (β) error probabilities set at 0.05 and 0.2, respectively[16]. TSA was applied to the following five primary outcome measures:

1. Return of Spontaneous Circulation (ROSC)
2. Time to Airway Securement
3. First-pass Success Rate of Airway Insertion
4. Favorable Neurological Outcome
5. Occurrence of Regurgitation/Aspiration

### 2.5 Heterogeneity analysis

We assessed the heterogeneity of the combined data using the following steps: First, we used an I^2^ measure to assess whether there was any heterogeneity between the combined literature. I^2^ ≥ 75% showed high heterogeneity, 50% ≤ I^2^ < 75% showed moderate heterogeneity, and 25 ≤ I^2^ < 50% showed low heterogeneity. If I^2^ = 0, we used the fixed-effect model for data analysis; otherwise, the random-effect model was used. If there was any heterogeneity among the combined data, we conducted a sensitivity analysis or subgroup analysis to analyze the source of heterogeneity.

## 3 Results

We identified 4,158 articles. Among these, 1,048 were duplicates. By reviewing titles and abstracts, we excluded 3,023 studies unrelated to our research. During full-text screening, we excluded 18 articles due to flawed or unclear study design, 24 articles with incorrect interventions, 6 articles reporting outcomes unrelated to our predefined endpoints, and 7 articles with missing data. Ultimately, 9 randomized controlled trials (RCTs) and 23 non-randomized controlled studies met the inclusion criteria and were incorporated into the meta-analysis. The literature search and study selection process are illustrated in the flowchart presented in Figure 1

**Figure 1.**
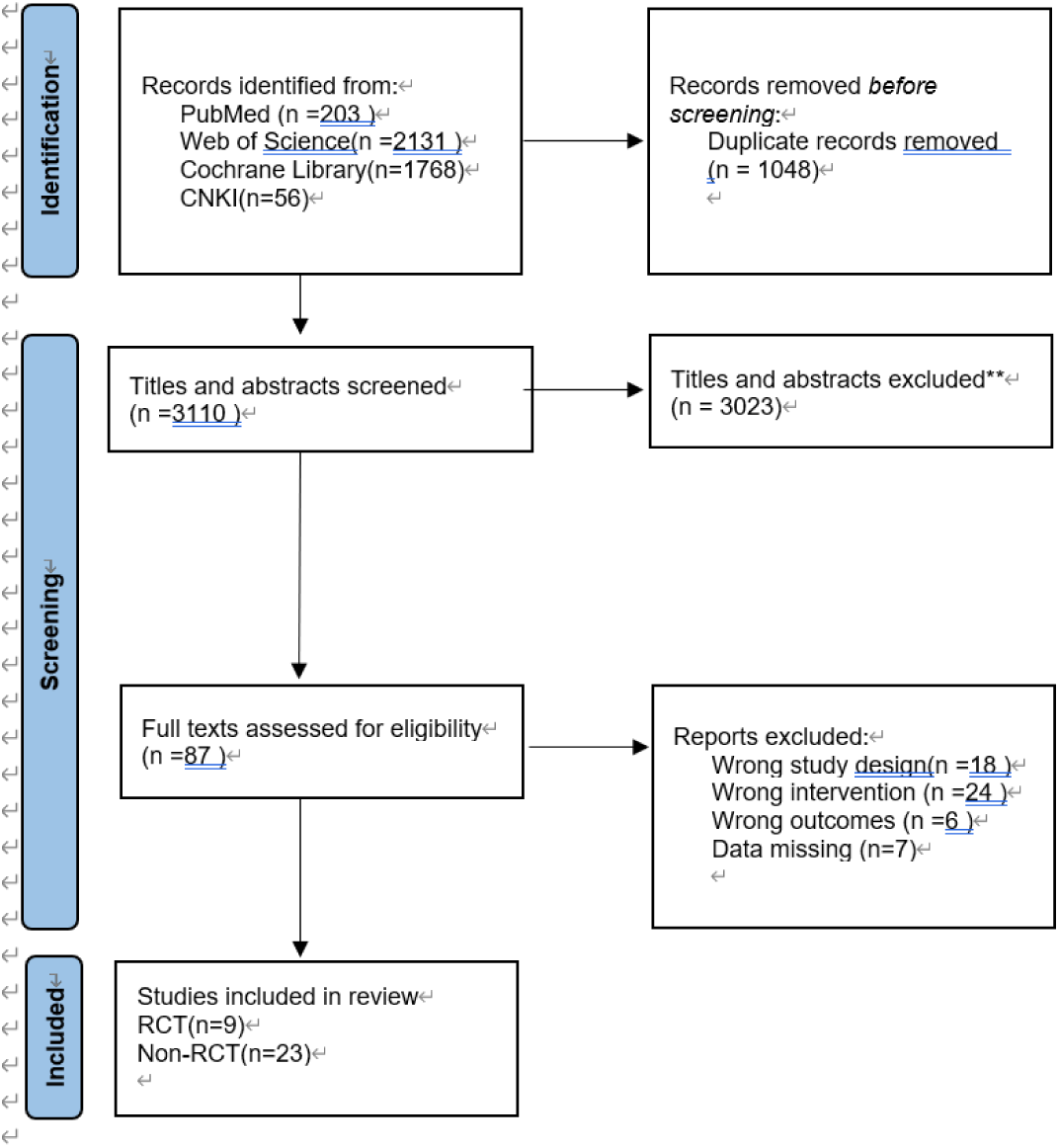
The process of literature retrieval and study selection.

### 3.1 Description of Included Studies

A total of 32 studies were included in this meta-analysis: 9 randomized controlled trials (RCTs) and 23 non-randomized controlled studies (non-RCTs). All studies enrolled patients experiencing out-of-hospital cardiac arrest (OHCA) requiring advanced airway management.

Regarding outcome reporting:

- Return of Spontaneous Circulation (ROSC) was reported in 7 RCTs [8, 12, 17–21]and 17 non-RCTs.[11, 22–37]
- First-pass success rate of advanced airway insertion was reported in 7 RCTs[12, 17–21, 38] and 9 non-RCTs.[11, 23–25, 31, 39–42]
- Time required for advanced airway insertion was reported in 7 RCTs[12, 18–21, 38, 43] and 6 non-RCTs.[11, 23, 25, 31, 32, 34]
- Neurological outcomes were reported in 3 RCTs[8, 12, 18] and 9

non-RCTs.[22, 26, 27, 33–35, 37, 44, 45]

- Regurgitation/aspiration was reported in 4 RCTs.[8, 17, 18, 21] Concerning the study populations:
- Among the 9 RCTs, 5 enrolled Asian populations and 4 enrolled Caucasian populations.
- Among the 23 non-RCTs, 10 enrolled Asian populations and 13 enrolled Caucasian populations.

The results of the risk of bias assessments using the ROB2 tool for RCTs and the ROBINS-I tool for non-RCTs are presented in the figure 2 below.

**Figure 2:**
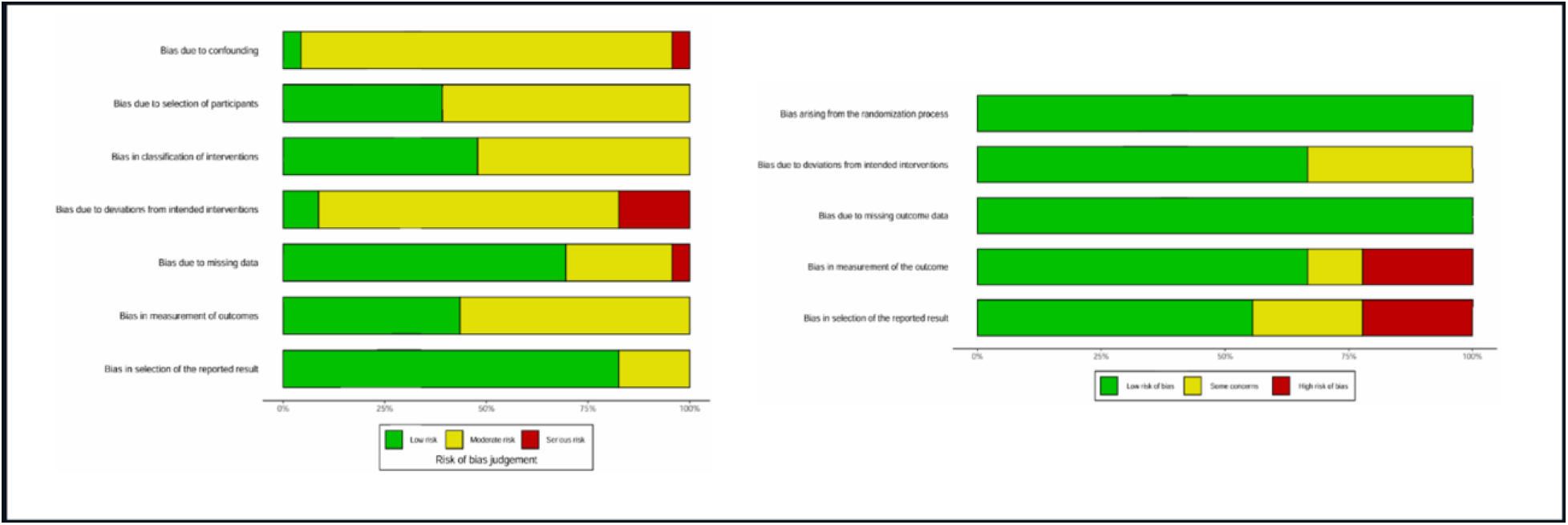
The results of the risk of bias assessments

### 3.2 ROSC

This study included a total of 24 studies that evaluated the impact of supraglottic airway devices (SGA) versus endotracheal intubation (ETI) on the return of spontaneous circulation (ROSC) in adult out-of-hospital cardiac arrest (OHCA) patients. Among these, seven were randomized controlled trials (RCTs) and 17 were non-randomized studies (non-RCTs). Meta-analyses and subgroup analyses were conducted separately for these two study types.

In the seven RCTs, the pooled results demonstrated a significantly higher ROSC rate in the SGA group compared to the ETI group (RR = 1.17, 95% CI: 1.03–1.34, *P* = 0.016), with moderate heterogeneity (I² = 48.5%). In the Caucasian subgroup, the pooled RR was 1.09 (95% CI: 1.03–1.17), with very low heterogeneity (I² = 0%), indicating stable results. In the Asian subgroup, the pooled RR was 1.39 (95% CI: 0.97–1.99); this result was not statistically significant, and heterogeneity was high (I² = 69.6%). No statistically significant difference in effect was observed between these two subgroups (*P* = 0.1992), suggesting that ethnicity may not be the primary source of the differential intervention effects (Figure 3A).

**Figure 3:**
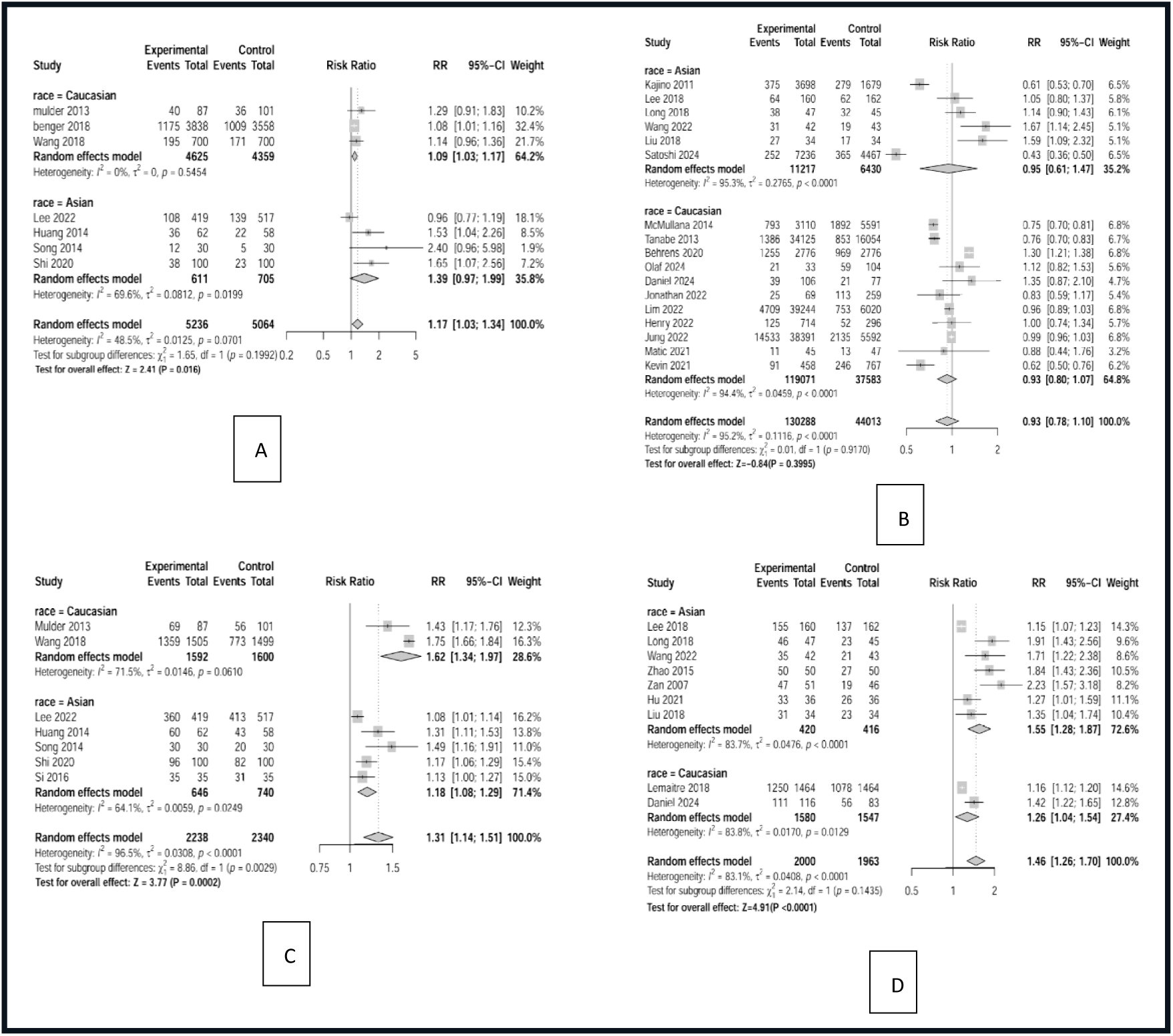
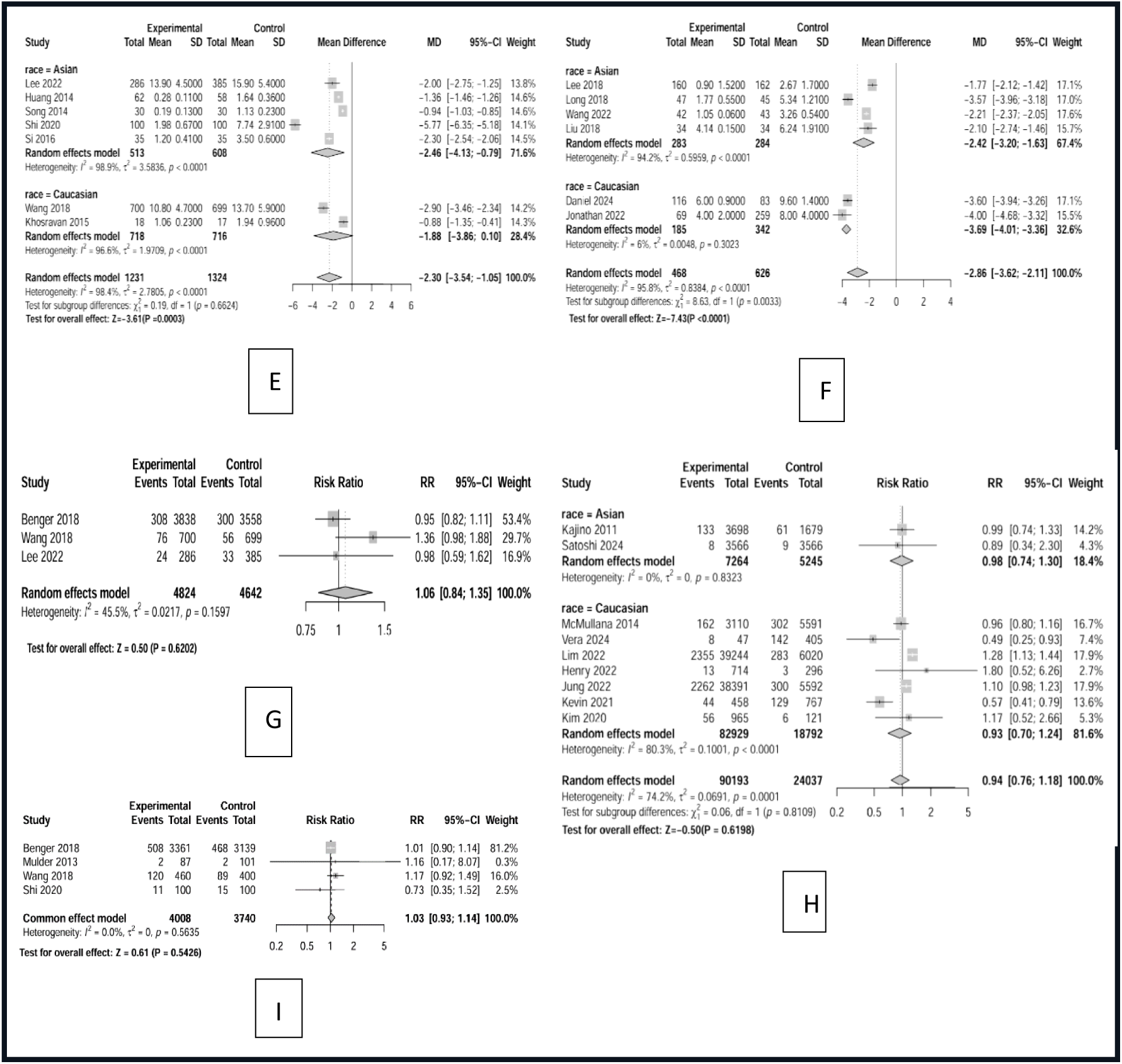
The result of forest plot: (A:ROSC in RCT, B:ROSC in non-RCT, C: first-attempt success rates in RCT, D: first-attempt success rates in non-RCT, E: Time to Airway Securement in RCT, F: Time to Airway Securement in non-RCT, G: Favorable Neurological Outcome in RCT, H: Favorable Neurological Outcome in non-RCT, I: Occurrence of Regurgitation/Aspiration)

In the 17 non-randomized studies, the overall pooled analysis showed no statistically significant difference in ROSC rates between the SGA and ETI groups (RR = 0.93, 95% CI: 0.78–1.10, *P* = 0.3995). However, heterogeneity was extremely high (I² = 95.2%, *P* < 0.0001). In the Asian subgroup, the pooled RR was 0.95 (95% CI: 0.61–1.47), with considerable heterogeneity (I² = 95.3%). In the Caucasian subgroup, the RR was 0.93 (95% CI: 0.80–1.07), also exhibiting extreme heterogeneity (I² = 94.4%). No significant difference was found between subgroups (*P* = 0.9170). Overall, the non-randomized studies suggest no significant difference in ROSC between SGA and ETI; however, due to the high degree of inconsistency across studies, these results should be interpreted with caution (Figure 3B).

### 3.3 first-attempt success rates

The study included 16 studies evaluating the impact of supraglottic airway devices (SGA) versus endotracheal intubation (ETI) on first-attempt success rates in adult out-of-hospital cardiac arrest (OHCA) patients. Among these, 7 were randomized controlled trials (RCTs), and 9 were non-randomized controlled studies (non-RCTs). Meta-analyses and subgroup analyses were conducted separately.

In the 7 RCTs, the pooled results showed that the SGA group had a significantly higher first-attempt success rate than the ETI group (RR = 1.31, 95% CI: 1.14–1.51, P = 0.0029), with considerable heterogeneity (I² = 96.5%). Subgroup analysis by ethnicity revealed that in the Caucasian subgroup, the pooled RR was 1.62 (95% CI: 1.34–1.97), with moderate heterogeneity (I² = 71.5%). In the Asian subgroup, the pooled RR was 1.18 (95% CI: 1.08–1.29), also with moderate heterogeneity (I² = 64.1%). After subgroup analysis, heterogeneity decreased compared to the overall pooled analysis, and the difference in effects between subgroups was statistically significant (P = 0.0029), suggesting that ethnicity may be a major source of variation in intervention effects. (Figure 3C) In the 9 non-RCTs, the overall pooled analysis showed no statistically significant difference in first-attempt success rates between SGA and ETI (RR = 1.46, 95% CI: 1.26–1.70, P < 0.0001), but with high heterogeneity (I² = 83.1%, P < 0.0001). In subgroup analyses, the Asian subgroup had a pooled RR of 1.55 (95% CI: 1.28–1.87), with considerable heterogeneity (I² = 83.7%), while the Caucasian subgroup had an RR of 1.26 (95% CI: 1.04–1.54), also with high heterogeneity (I² = 83.8%). The difference between subgroups was not statistically significant (P = 0.1435). Overall, the non-randomized studies suggested a significant difference in first-attempt success rates favoring SGA over ETI, but interpretations should be made with caution due to high heterogeneity. (Figure 3D)

### 3.4 Time to Airway Securement

We included 13 studies evaluating the impact of supraglottic airway devices (SGA) versus endotracheal intubation (ETI) on airway insertion time in adult out-of-hospital cardiac arrest (OHCA) patients. Among these, 7 were randomized controlled trials (RCTs), and 6 were non-randomized controlled studies (non-RCTs). Meta-analyses and subgroup analyses were conducted separately.

In the 7 RCTs, pooled results showed that the airway insertion time in the SGA group was significantly shorter than in the ETI group (MD = -2.30 min, 95% CI: -3.54 to -1.05, P = 0.0003), with considerable heterogeneity (I² = 98.4%). Subgroup analysis by ethnicity revealed that in the Caucasian subgroup, the pooled MD was -1.88 min (95% CI: -3.86 to 0.10), with considerable heterogeneity (I² = 96.6%). In the Asian subgroup, the pooled MD was -2.46 min (95% CI: -4.13 to -0.79), also with considerable heterogeneity (I² = 98.9%). The difference in effects between subgroups was not statistically significant (P = 0.6624), suggesting that ethnicity may not be a major source of variation in intervention effects. (Figure 3E)

In the 6 non-RCTs, the overall pooled analysis showed a statistically significant difference in airway insertion time between SGA and ETI (MD = -2.86 min, 95% CI: -3.62 to -2.11, P < 0.0001), but with high heterogeneity (I² = 95.8%). In subgroup analyses, the Asian subgroup had a pooled MD of -2.42 min (95% CI: -3.20 to -1.63), with considerable heterogeneity (I² = 94.2%). The Caucasian subgroup had an MD of -3.69 min (95% CI: -4.01 to -3.36), with very low heterogeneity (I² = 6%). The difference between subgroups was statistically significant (P = 0.0033), suggesting that ethnicity may be a major source of heterogeneity. Overall, non-randomized studies indicated a significant difference in airway insertion time favoring SGA over ETI. (Figure 3F)

### 3.5 Favorable Neurological Outcome

We included 12 studies evaluating the impact of supraglottic airway devices (SGA) versus endotracheal intubation (ETI) on neurological outcomes in adult out-of-hospital cardiac arrest (OHCA) patients. Among these, 3were randomized controlled trials (RCTs), and 9 were non-randomized controlled studies (non-RCTs). Meta-analyses and subgroup analyses were conducted separately.

In the 3 RCTs, pooled results showed no significant difference in favorable neurological outcomes between the SGA group and the ETI group (RR = 1.06, 95% CI: 0.84–1.35, P = 0.6202), with low heterogeneity (I² = 45.4%). (Figure 3G)

In the 9 non-RCTs, the overall pooled analysis showed no statistically significant difference in the rate of favorable neurological outcomes between SGA and ETI (RR = 0.94, 95% CI: 0.76–1.18, P = 0.6198), with moderate heterogeneity (I² = 74.2%). Subgroup analyses revealed that in the Asian subgroup, the pooled RR was 0.98 (95% CI: 0.74–1.30), with very low heterogeneity (I² = 0%). In the Caucasian subgroup, the RR was 0.93 (95% CI: 0.70–1.24), with high heterogeneity (I² = 80.3%). The difference between subgroups was not statistically significant (P = 0.1597). Overall, the results suggest no significant difference in neurological outcomes between SGA and ETI. (Figure 3H)

### 3.6 Occurrence of Regurgitation/Aspiration

Only 4 of the 32 included randomized controlled trials reported regurgitation/aspiration events. Compared with tracheal intubation, the use of SGAs (Supraglottic Airways) may have no effect on regurgitation/aspiration events (RR 1.03; 95% CI, 0.93-1.14; P=0.5426), with very low heterogeneity, indicating robust results. (Figure 3I

We did not pool data on airway displacement because reporting and definitions were inconsistent across the included trials. In one study, unrecognized airway misplacement/displacement occurred in 10 of 1,353 patients (0.7%) assigned to SGA and tracheal displacement occurred in 24 of 1,299 patients (1.8%) assigned to tracheal intubation.

1. [18] .Whereas in another trial, unintended loss of the previously established airway occurred in 412 of 3,900 patients (10.6%) assigned to SGA and in 153 of 3,081 patients (5.0%) assigned to tracheal intubation.[8] Finally, we also conducted a GRADE evidence grading to explore the level of this evidence. The Certainty of evidence is moderate or high. The detailed results are provided in the additional materials.(Supplementary materials 1)

### 3.7 Bias Assessment

Most outcomes are in low bias. (Supplementary materials 2)

### 3.8 Sensitivity Analysis

The results of the analysis were robust to both post hoc sensitivity analyses with similar results and conclusions. (Supplementary materials 2)

### 3.9 Trial Sequential Analysis (TSA)

Finally, we applied TSA analysis, and the results showed that, apart from the outcome indicators of good neurological prognosis and aspiration, the TSA results for the remaining outcome indicators, although not reaching the RIS, still had cumulative Z-curves that crossed the conventional and monitoring boundaries, indicating firm evidence of the superiority of supraglottic airway devices (SGA) over endotracheal intubation (ETI) in OHCA patients before reaching the RIS (Supplementary materials 2).

## 4 DISCUSSION

According to this study, to use supraglottic airway (SGA) as the first method for prehospital airway management results in faster placement, greater success on the first attempt, and higher rates of return of spontaneous circulation (ROSC) when compared to endotracheal intubation (ETI).

In prehospital airway management, regardless of the device used, rapid placement and ensuring effective ventilation are critical for out-of-hospital cardiac arrest (OHCA) patients. Prompt airway placement and effective ventilation are key to successful resuscitation and preventing hypoxic brain injury. [46]In this study, across both randomized controlled trials (RCTs) and non-randomized studies, SGA placement time, first-pass success rate, and ROSC were either superior or non-inferior to ETI. Furthermore, some studies indicated no statistically significant differences in oxygen saturation, partial pressure of oxygen, or oxygenation index between patients managed with SGAs and those intubated.[21, 30, 38, 41, 47] This demonstrates equivalent ventilation efficacy between SGAs and ETI devices. For OHCA patients, SGA management may offer advantages or at least be non-inferior to ETI.

This study also shows that SGAs are equally as good as ETI regarding the complication of regurgitation and aspiration. The distal cuff of second-generation SGAs like the laryngeal mask airway (LMA) and i-gel seals the upper esophagus and surrounds the glottic opening. From a physical and ergonomic perspective, this design essentially eliminates the risk of aspiration. If esophageal regurgitation occurs, these SGAs can prevent gastric contents from entering the airway. [48]A large study involving 2,004 patients showed an aspiration rate of only 0.05% with the i-gel. Thus, SGAs can effectively prevent aspiration pneumonia, providing a better foundation for subsequent in-hospital care.[49] This study indicates SGAs are as effective as ETI in preventing regurgitation and aspiration complications.

Regarding neurological outcomes, SGAs also did not demonstrate inferiority. Although in the sequential trial analysis the Z-curve did not cross the conventional boundary or the trial sequential analysis (TSA) curve, potentially indicating the need for further exploration, we believe favorable neurological prognosis is also critically dependent on effective post-admission care. In cases like traumatic brain injury or severe hemorrhage leading to inadequate cerebral perfusion, the nervous system may already have sustained organic damage, limiting the impact of prehospital airway management on neurological outcomes at discharge[50].Crucially, this study did not show that using SGAs for OHCA airway management leads to worse neurological outcomes.

Significant heterogeneity was observed for some outcomes in this study, such as placement time and first-pass success in RCTs, and ROSC, placement time, favorable neurological outcome, and first-pass success in non-randomized studies. We hypothesize that the ethnicity of the study populations and the types of SGAs used may be major sources of this heterogeneity. Among the 9 included RCTs, 4 studied Caucasian populations and 5 studied Asian populations. Among the 23 included non-randomized studies, 13 involved Caucasian populations and 10 involved Asian populations. Subgroup analysis by ethnicity reduced heterogeneity in both groups, but some remained. This could be due to differences in glottic size and the proportion of difficult airways between Asian and Caucasian populations[51]. Additionally, while all included studies focused on OHCA patients, the causes of arrest varied. Variations in the healthcare levels and treatment practices across the countries/regions where the studies were conducted may also contribute. Furthermore, not all included studies, particularly the non-randomized ones, reported the specific types of SGAs used, which could be another source of heterogeneity. For the outcome "placement time," the definition varied significantly between studies. Some measured from arrival at the scene until airway management completion, while others measured only the duration from device insertion attempt start to successful placement. This inconsistency in defining the start and end points contributed to heterogeneity. Moreover, as we included retrospective and prospective cohort studies, it was challenging for researchers to fully control confounding factors, introducing a risk of information bias.

The results of our meta-analysis reversed previous results from meta-analysis of observational data, which suggested higher ROSC rate with ETI compared to SGA. [52, 53] This discrepancy can be explained by "resuscitation time bias": patients in whom ETI failed likely received an SGA later in the resuscitation attempt, and could reasonably be expected to have worse outcomes.[54] Our findings align more closely with recent systematic reviews and network meta-analyses of RCTs and quasi-randomized studies, and a systematic review and meta-analysis of RCTs, which found increased ROSC with SGAs compared to ETI.[13, 14]

This meta-analysis provides more robust and precise results than previous ones. First, we separately analyzed RCTs and non-randomized studies. The results from RCTs and non-randomized studies were consistent. Second, we used the RoB 2 tool to assess the certainty of evidence for RCTs and the ROBINS-I tool for non-randomized studies, enhancing confidence in the quality assessment. Third, we included additional outcomes potentially important to patients, clinicians, and healthcare decision-makers, such as time required for airway placement and first-pass success rate, which were not consistently reported before. Fourth, we included the study by Lee et al.[12] and the latest RCTs and non-randomized studies from different regions, increasing the information size, precision, and certainty of the results.

## 5 CONCLUSION

In adult patients with out-of-hospital cardiac arrest (OHCA), initial airway management using a supraglottic airway device (SGA), compared to endotracheal intubation, will lead to a higher rate of return of spontaneous circulation (ROSC), faster airway placement times, and higher first-pass success rates. Furthermore, SGA have no negative effect on long-term survival outcomes or aspiration events.

## Data Availability

Not applicable

## Acknowledgements

The authors appreciate Dr. Charles Damien Lu for his kind help with the English proofreading and editing.

## Author contributions

Li-fu Zheng was the lead author of the Cochrane review, the data on which this analysis was based. Li-fu Zheng conceived and delineated the hypotheses, designed the study, acquired, and analyzed the data, and wrote and edited the manuscript of the previous analysis. Li-fu Zheng edited the manuscript of the present analysis. Ming-wei Sun, and Lei Deng developed the original concepts for this systematic review and meta-analysis. Li-fu Zheng and Ying-yi Ding contributed to the screening of the eligible studies, data extraction, and data synthesis. Ying-yi Ding and Jin-zhou Feng contributed to the quality assessment of literatures. Li-fu Zheng and Jin-zhou Feng drafted the first version of this manuscript. All authors read and approved the final manuscript and take responsibility for its publication.

## Declarations

### Ethics approval and consent to participate

Not applicable.

### Consent for publication

Not applicable.

### Availability of data and materials

Not applicable.

